# Neural Dysregulation in Post-COVID Fatigue

**DOI:** 10.1101/2022.02.18.22271040

**Authors:** Anne M.E. Baker, Natalie J. Maffitt, Alessandro Del Vecchio, Katherine M. McKeating, Mark R. Baker, Stuart N. Baker, Demetris S. Soteropoulos

**Affiliations:** Faculty of Medical Sciences, Newcastle University, UK; Department Artificial Intelligence in Biomedical Engineering, Friedrich–Alexander University Erlangen–Nürnberg, Germany

**Keywords:** Fatigue, COVID, transcranial magnetic brain stimulation, myopathy, dysautonomia

## Abstract

Following infection from SARS-CoV-2, a substantial minority of people develop lingering after-effects known as ‘long COVID’. Fatigue is a common complaint with substantial impact on daily life, but the neural mechanisms behind post-COVID fatigue remain unclear.

We recruited 37 volunteers with self-reported fatigue after a mild COVID infection and carried out a battery of behavioural and neurophysiological tests assessing the central, peripheral and autonomic nervous systems.

In comparison to age and sex matched volunteers without fatigue (n=52), we show underactivity in specific cortical circuits, dysregulation of autonomic function, and myopathic change in skeletal muscle. Cluster analysis revealed no sub-groupings, suggesting post-COVID fatigue is a single entity with individual variation, rather than a small number of distinct syndromes. Based on our analysis we were also able to exclude dysregulation in sensory feedback circuits and descending neuromodulatory control.

These abnormalities on objective tests may indicate novel avenues for principled therapeutic intervention, and could act as fast and reliable biomarkers for diagnosing and monitoring the progression of fatigue over time.

## Introduction

Most people infected with SARS-CoV-2 do not require hospitalisation. However, even after a mild infection, a minority develop symptoms that linger for weeks or months (long COVID). Persistent fatigue, where everyday actions become laborious, is one of the more commonly reported after-effects^1^ and can have a substantial impact on the quality of life and productivity of sufferers^2-4^. At the time of publication ∼2% of the UK population are experiencing long COVID; >50% report fatigue as their primary symptom^5^.

Fatigue appears to be a multi-system pathology associated with immunological, metabolic and hormonal anomalies. There are strong links between the immune and nervous systems with multiple pathways for possible interactions^6^. These presumably generate changes in neurological function, which in turn lead to feelings of weakness, with physical and cognitive actions being more effortful. Such effects could result from changes at many levels of the nervous system; here we focused on five potential neural substrates of fatigue, that might not only result in increased performance fatigue but also increased perception of fatigue ^7^:

### Hypothesis 1

Motoneurons (and the muscles they innervate) are activated by multiple inputs from motor cortical areas, the brainstem and spinal cord. If any of these systems have reduced excitability or increased inhibition, as demonstrated in other chronic conditions associated with fatigue^8^, this could contribute to a perception of fatigue.

### Hypothesis 2

During normal self-generated movements sensory feedback is attenuated^9^. Incomplete sensory attenuation during movement could lead to heightened feedback, and an increase sense of effort^10^.

### Hypothesis 3

At the level of the periphery, other post-viral syndromes (such as Guillain-Barré and Miller Fisher syndrome) often lead to ineffective signal transmission at the neuromuscular junction, whereas myopathic changes within the muscle fibres themselves will cause weakness^11-13^ requiring stronger voluntary drive to generate force, which could give rise to an increased perception of effort^14^.

### Hypothesis 4

Monoaminergic neuromodulators are released in the spinal cord and regulate the gain of motoneuron responses to inputs through the activation of specific membrane conductances^15^. If neuromodulatory inputs to motoneurons are affected after post-COVID fatigue (pCF)^16^, a stronger synaptic drive would be required for a given level of force. This could contribute to movements being perceived as more effortful.

### Hypothesis 5

Autonomic dysregulation is often a predictor for fatigue in other chronic illnesses^17^, and treating dysautonomia has shown promising results in the improving the symptoms of fatigue^18^. Autonomic dysregulation could also contribute to pCF.

In this study, we used an extensive battery of non-invasive tests to compare pCF sufferers with a matched control group, testing these varied hypotheses. Our results provide evidence for some of the hypothesised mechanisms, while suggesting that others are unlikely to contribute. pCF seems to result from dysregulation in specific components of the central, peripheral and autonomic nervous systems.

## Methods

To understand the neural mechanisms behind pCF, we utilised a wide range of well-characterised non-invasive behavioural and neurological tests (summarized in Fig. 1 and described in detail in the Supplementary Information). Through these tests we were able to probe specific components within the central (CNS), peripheral (PNS) and autonomic (ANS) nervous systems. Transcranial magnetic stimulation (TMS) probed the state of intracortical motor circuits. Sensory nerve stimulation assessed the impact of sensory feedback on the central nervous system. Electrical stimulation of muscles assessed both central and peripheral levels of fatigue, while recordings of heart rate and galvanic skin responses assessed the state of the autonomic nervous system. High density surface electromyography extracted the activity of muscle motor units, from which we derived metrics of the state of neuromodulatory systems. Collectively these tests yielded 35 measures (33 relating directly to the state of the nervous system, plus blood oxygen saturation and tympanic temperature). These measures are referred to using consistent abbreviations in this report, as defined in the Supplementary Information; in the text these abbreviations are in italics. Participants also completed a Fatigue Impact Scale (FIS)^19^ questionnaire via a web-based survey tool.

**Figure 1.**
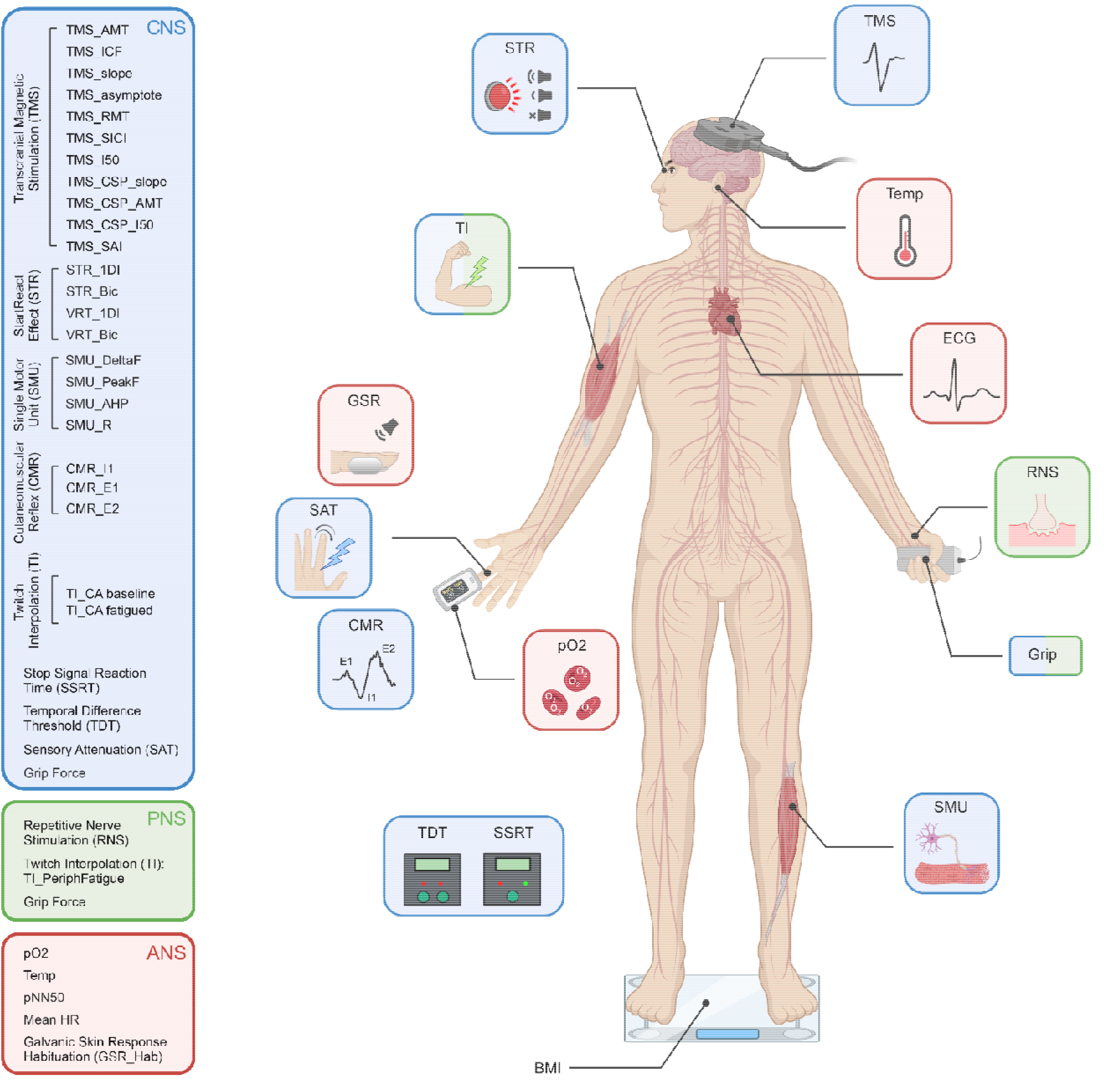
Schematic representation of the different tests performed, colour coded according to which components of the central, peripheral and autonomic nervous systems (CNS, PNS, ANS) they assessed. TMS, transcranial magnetic stimulation; ECG, electrocardiogram; RNS, repetitive nerve stimulation; SMU, single motor unit recording; BMI, body mass index; TDT, temporal difference threshold; SSRT, stop signal reaction time; pO2, blood oxygen saturation; CMR, cutaneomuscular reflex; SAT, sensory attenuation with movement; GSR, habituation of the galvanic skin response to loud sound; TI, twitch interpolation, STR, StartReact effect. Created with biorender.com.

Tests were carried out on two groups of volunteers - one who were self-reporting as suffering from pCF, and a second cohort of control subjects with no fatigue. Inclusion criteria were age 18-65 years with no history of neurological disease and 6-26 weeks after infection (for the pCF cohort). The study was approved by the Ethics Committee of the Newcastle University Faculty of Medical Sciences; participants provided written informed consent to take part.

### Statistical Methods

Descriptive statistics are given as mean±SD. Each of the measures we collected had different units and scales. To allow easy comparison of differences between measures, and to avoid a metric with large values dominating the classification algorithm (see below), data were normalised as a Z score for each feature. This was computed by finding the difference between the mean of a measure between the pCF and control cohorts, and dividing by the standard deviation of the control cohort. To correct for multiple comparisons, we used the Benjamini-Hochberg procedure^20^. Raw (uncorrected) P values are given throughout this report, together with a statement of whether these values passed the Benjamini-Hochberg procedure.

### Data Availability

A spreadsheet containing Z-transformed values for measurements in all subjects is available in the Supplementary Information.

## Results

A total of 39 people with pCF, and 53 controls who were not suffering from pCF were initially recruited to the study. Prior to attending the laboratory, volunteers with pCF underwent a structured telephone interview, which checked details of their medical history and possible exclusion criteria. Further measurements were then made during a single laboratory visit lasting around four hours. Two participants with pCF were discovered during the course of the study to be under clinical investigation for neurological symptoms and signs not part of the typical long COVID syndrome. One additional participant from the control group was found to have an exaggerated startle response even to weak stimuli, which precluded gathering meaningful data on many of the protocols. These three individuals were excluded from the database, leaving 37 pCF (27 female, 73%) and 52 controls (37 female, 71%). The two cohorts were well matched for age, as illustrated by the cumulative distribution plots in Fig. 2A (and were not significantly different, p>0.5, unpaired t-test). Full demographic information about the two cohorts is available in Supplementary Table 1.

**Figure 2.**
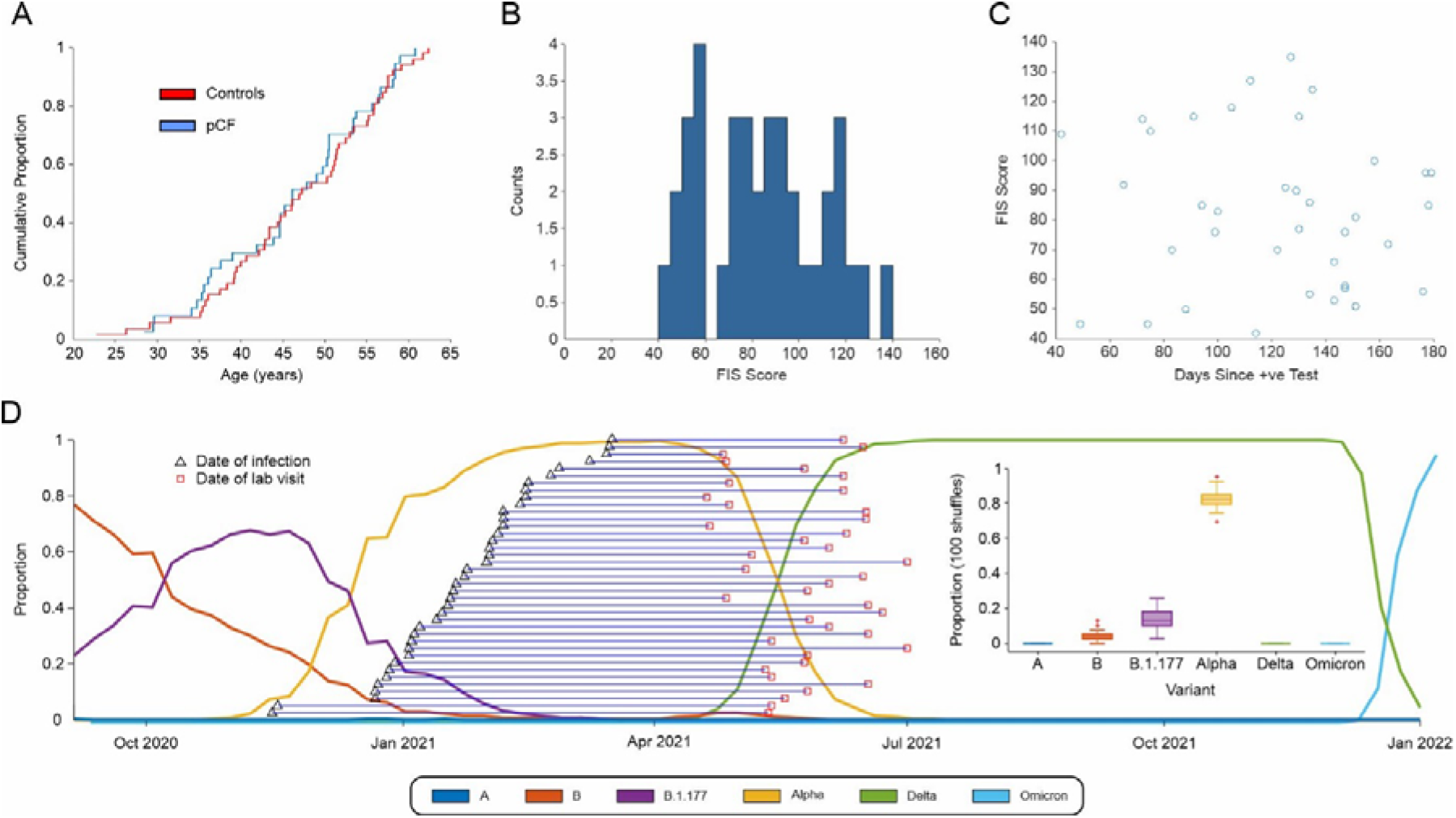
A, cumulative age distribution plots for pCF and control subjects. B, distribution histogram of FIS scores reported by pCF subjects. C, lack of correlation of FIS score with time since SARS-CoV-2 infection (r^2^=0.009, P=0.59). D, proportions of the most common SARS-CoV-2 variants in circulation in England since October 2020 and the estimated expected proportion of each variant across our cohort.

The FIS score reflects functional limitation due to fatigue experienced within the last 4 weeks, rather than a measure of the level of fatigue, and for the pCF cohort the mean score was 83±26 (range 42-135; Fig. 2B), suggesting, on average, a moderate impact in daily life. The interval between diagnosis with SARS-CoV-2 and attending the laboratory was 121±37 days (range 42-179 days). There was no correlation between the severity of fatigue measured by FIS score and time since infection (Fig. 2C; r^2^=0.009, P=0.59).

Although we do not have any way of definitively knowing the virus variant that our fatigue participants were infected with, we can estimate the likely proportions based on the known distribution of variants at the time. The weekly proportion of the six main variants circulating in England since Nov 2020 (A, Alpha, B, B.1.177, Delta & Omicron) was downloaded from the Sanger Institute COVID 19 Genomic Surveillance website (https://covid19.sanger.ac.uk/lineages/raw). For each subject in the pCF cohort we randomly assigned a variant 100 times, with a probability based on the relative proportions of variants at the time of their week of infection. By collating all the data across all pCF subjects, we could then estimate the expected proportions of each variant across our fatigue cohort (shown in Fig. 2D). Based on the published relative incidence of SARS-CoV-2 variants in the UK, we thus estimate 83%±5% of our pCF cohort had the Alpha variant.

Figure 3A presents the normalised data for each metric as a spider plot^21^, ordered so the greatest difference is located at the top of the figure; the shading indicates the standard error of the mean difference (calculated by dividing the standard deviation of each metric by the square root of number of data points available). The significance of differences between the pCF and control cohorts was assessed using unpaired t-tests. We highlight the ten measures which had uncorrected P<0.05 with coloured boxes on Fig. 3A; Fig. 3B compares the distribution of these measures between the cohorts as box-and-whisker plots. Four of the measures had differences so great, that they were assessed as significantly different even after adjustment for multiple comparisons; these are indicated with thicker lines in Fig. 3A. Full descriptive statistics for all measures in both cohorts are given in Table 1.

**Figure 3.**
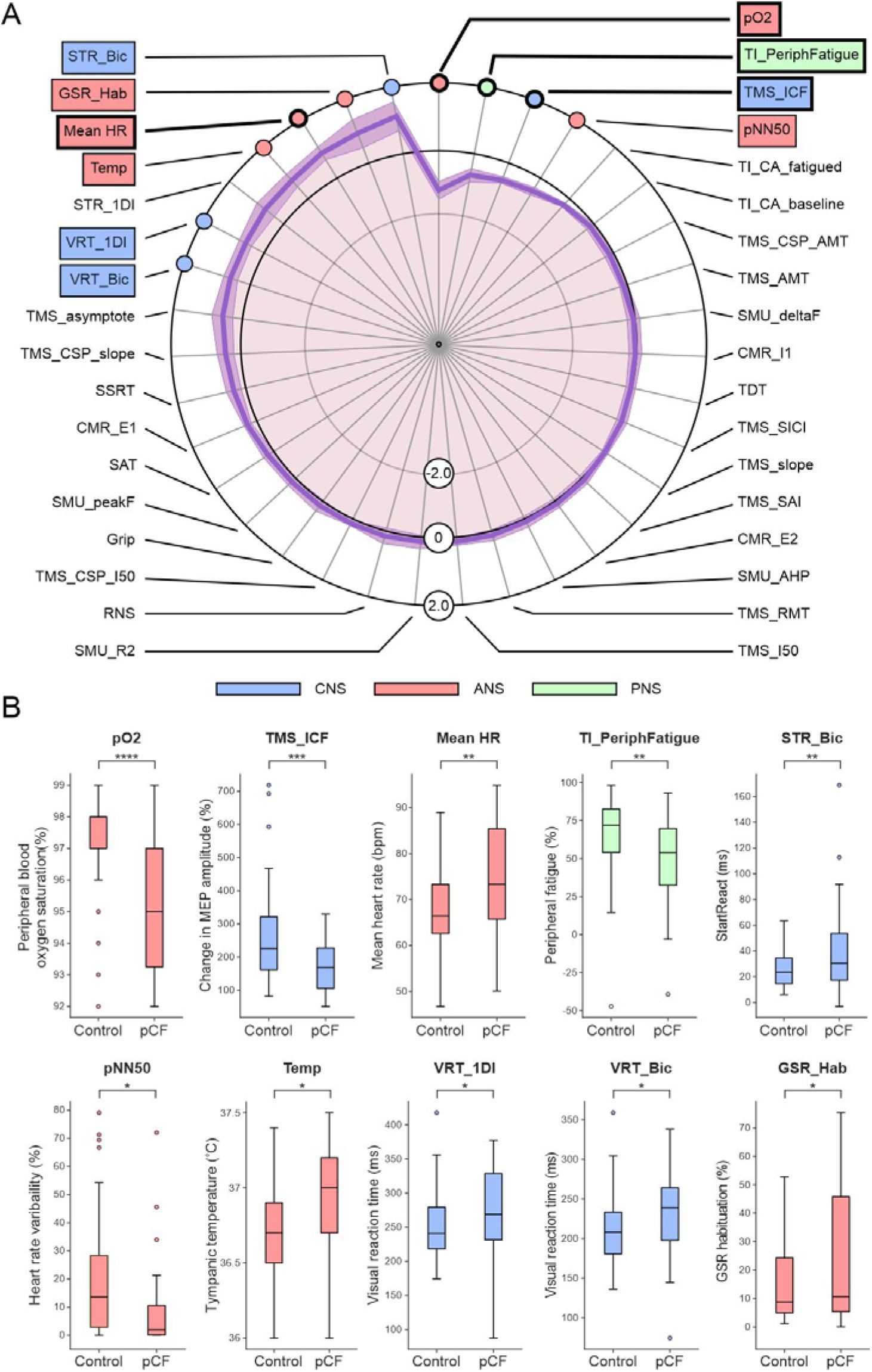
A, results from the tests outlined in Fig. 1, normalised as Z scores (difference between pCF and control subjects, scaled by SD). Measures highlighted with coloured boxes were individually significantly different between pCF and controls (P<0.05); for those with thicker lines, significance passed the Benjamini-Hochberg correction for multiple comparisons. B, distribution of the ten measures which had uncorrected P<0.05 as box-and-whisker plots across the two cohorts.

Voluntary activation of muscles relies on command signals from motor areas of the cortex; the state of cortical circuits has been linked to perception of effort and force output during fatiguing contractions^22,23^. By using transcranial magnetic stimulation (TMS) to assess the function of primary motor cortex we found that intracortical facilitation (*TMS_ICF*^*24*^) was significantly lower in pCF than controls (conditioned motor evoked potential relative to unconditioned 171±79% *vs* 258±140%, p<0.001), suggesting reduced cortical excitability (hypothesis 1). Other TMS measures also likely to be related to cortical excitability were no different between controls and pCF (the asymptote of the TMS recruitment curve, *TMS_asymptote*; the recruitment curve slope, *TMS_slope*; the intensity yielding 50% of the asymptote response amplitude, *TMS_I50*; active motor threshold, *TMS_AMT*; resting motor threshold, *TMS_RMT*). Multiple measures of cortical inhibition showed no significant differences between pCF and controls (short-interval intracortical inhibition, TMS_SICI; metrics of cortical silent period *TMS_CSP_AMT, TMS_CSP_slope, TMS_CSP_I50*). Possibly consistent with reduced cortical excitability, we also found a trend towards longer visual reaction times in pCF (in biceps muscle, *VRT_Bic*, 232±52ms *vs* 210±41ms; P=0.026; in first dorsal interosseous muscle, *VRT_1DI*, 277±61ms *vs* 251±46ms, P=0.024 for pCF *vs* controls respectively; neither P value crossed the significance threshold after adjustment for multiple comparisons).

Disturbances in sensory feedback processing have been previously hypothesised to contribute to an increased perception of effort^25^ (hypothesis 2). However, the attenuation of sensory input during movement (*SAT*), short-latency afferent inhibition (*TMS_SAI*) and the different components of the cutaneomuscular reflex (*CMR_E1, CMR_I1, CMR_E2*) all showed no significant differences. This suggests that sensory abnormalities are unlikely to be a contributing factor to pCF in our cohort.

Fatigue could arise from a reduced ability of the neuromuscular apparatus to generate force; a given movement would then require stronger voluntary drive and perceived effort would rise. Changes could arise in muscles themselves^26^, due to a weakened connection from motoneurons to muscles fibres^27^, or because motoneurons are less excitable. We found that maximal grip strength (*Grip*) was not significantly reduced in pCF, suggesting no deficit in force production for brief contractions. The efficacy of transmission at the neuromuscular junction (assessed using repetitive nerve stimulation, *RNS*), and intrinsic motoneuron excitability (assessed by estimating the peak firing rate of single motor units, *SMU_peakF* and the after-hyperpolarisation of motoneurons, *SMU_AHP*) were also not significantly different between our two cohorts. However, when we tested changes during a prolonged maximal contraction, we found pCF subjects had an increased level of peripheral fatigue (size of maximal electrically-evoked twitch after sustained contraction compared to baseline, *TI_PeriphFatigue*, 48.5±30.8% in pCF vs 67.1±25.2% in controls, p=0.003). This suggests that people with pCF develop metabolic changes in muscle fibres after prolonged activity leading to reduced force output (hypothesis 3).

We assessed the state of descending neuromodulatory pathways by looking at differences in the recruitment and de-recruitment of motoneurons (*SMU_deltaF*); the persistent inward currents that mediate this phenomenon are highly sensitive to serotonergic and noradrenergic inputs. We did not find any difference between our cohorts, suggesting that pCF is not associated with significant changes in descending neuromodulatory drive (hypothesis 4).

Autonomic dysregulation is often associated with fatigue in other conditions^17,28^ and recent studies reported autonomic dysregulation after SARS-CoV-2 infection^29-32^ (although not universally^33^). We found a significantly increased resting heart rate in pCF (*Mean_HR*, 74.8±11.1 vs 67.7±8.8 beats/min, p=0.0016). Other measures of autonomic function (tympanic temperature, *Temp*, 36.9±0.4°C *vs* 36.7±0.3°C, P=0.018; heart rate variability, *pNN50*, 8.8±15.7% *vs* 20.2±21.1%, P=0.011; galvanic skin response habituation, *GSR_Hab*, 25.2±24.5% *vs* 14.3±12.2%, P=0.026) also differed between cohorts, but did not pass correction for multiple comparisons. These results all point towards an increased vagal (relative to sympathetic) tone, suggesting pCF subjects suffer from a degree of dysautonomia (hypothesis 5).

Various behavioural measures did not show differences between pCF and control subjects. These included temporal difference threshold (*TDT*^*34*^) and stop signal reaction time (*SSRT*^*35*^); both are likely to be partly sensitive to inhibition in sub-cortical circuits. Central activation, which assesses the ability of the CNS to activate muscle maximally voluntarily, was also not different in pCF, either assessed at baseline (*TI_CA_baseline*) or after a fatiguing contraction (*TI_CA_fatigue*). The StartReact effect, which measures the acceleration of a visual reaction time by a loud (startling) sound and has been proposed to assess reticulospinal pathways, showed a trend to increase in the biceps muscle in pCF subjects (*STR_Bic*, 38.7±34.1ms vs 25.1±12.3ms, P=0.010) but did not survive correction for multiple comparisons and was not significantly different in the first dorsal interosseous (*STR_1DI*). This is likely to be driven by the increased visual reaction time in pCF described above; because the startle reaction times were similar to controls, this led to an elevated difference.

The level of common input to a motoneuron pool can assess cortical control of muscles; this was not different between pCF and controls (*SMU_R2*). Finally, we found a significant reduction in blood oxygen saturation in the pCF subjects (*pO2*, 95.3±1.9% vs 97.2±1.5%, P=0.00002).

Overall we found ten measures which were different between pCF and control subjects (uncorrected P<0.05), of which four passed significance correction for multiple comparisons. We investigated whether these dysregulations occurred in the same people, or whether pCF could be sub-divided into two or more different syndromes. K-means clustering followed by gap analysis revealed that the optimal cluster number was one (Fig. 4A), regardless of whether we included all metrics, or the four which were significantly different after correction for multiple comparisons. Dysregulations thus appear to vary independently across the pCF population, rather than being clustered in particular subsets.

**Figure 4.**
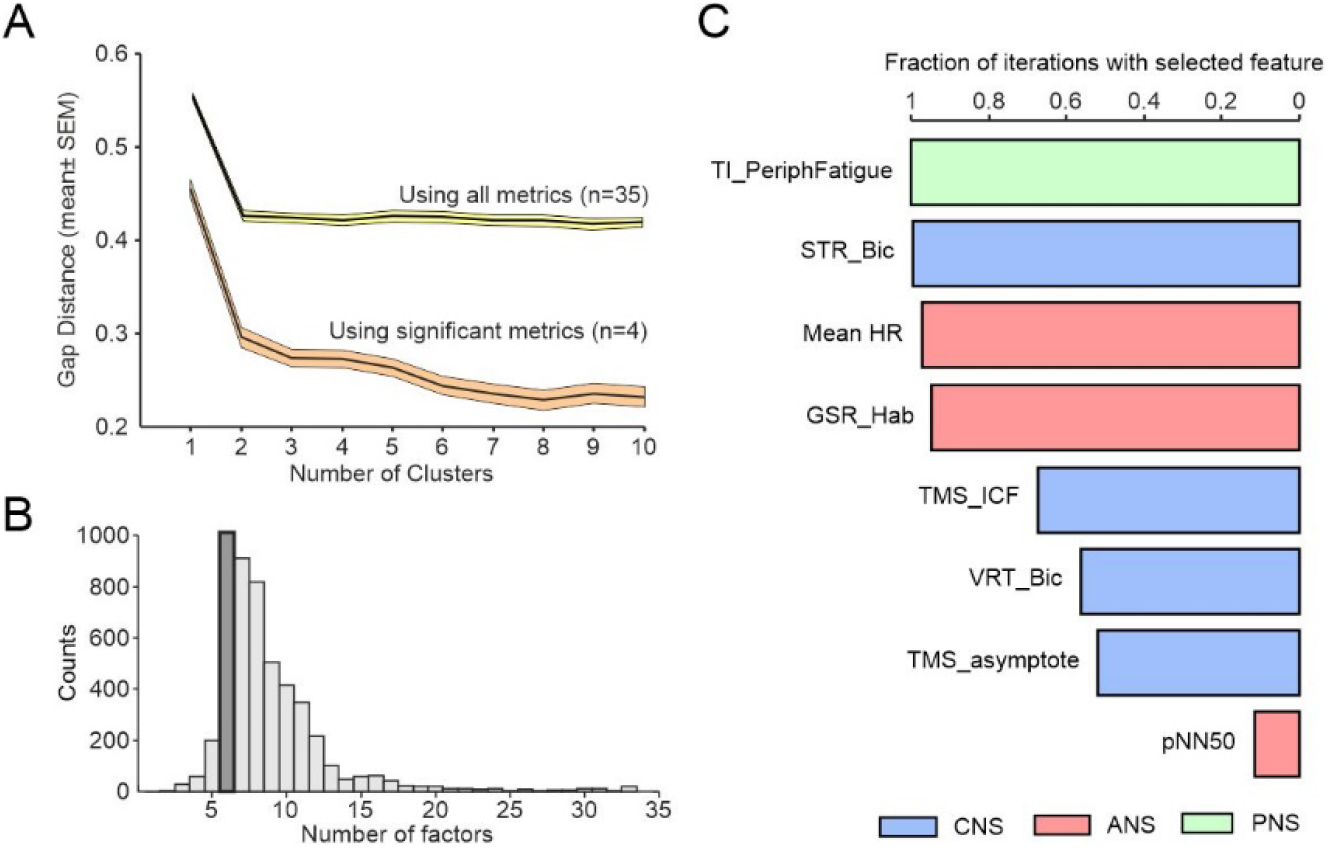
A, gap analysis of number of clusters in the multivariate dataset from pCF subjects. B, number of factors chosen by a machine learning algorithm to maximise classification of pCF *versus* control subjects. C, fraction of iterations of classification algorithm, with feature number fixed to six, which included different features. Plot has been truncated to show the most common eight features.

For the metrics which failed to reach significance individually, it was important to determine if they could still distinguish between pCF and controls in combination. We used a machine learning approach to classify participants as pCF or control, based on the multivariate data. Out of 33 available neurophysiological or behavioural metrics, repeated runs of the classifier determined the optimal feature number to maximise classifier accuracy. This had a mode of six (Fig. 4B). We then ran repeated classifications locked to six features, and counted how often a given measure was used (Fig. 4C). With this approach, the mean classification accuracy was 70% (SD 3.6%). In addition to the three neurophysiological features which were individually significantly different as described above (*TMS_ICF, Mean_HR, TI_PeriphFatigue*), additional frequently selected metrics were the habituation of the galvanic skin response to a startling stimulus and heart rate variability (*GSR_Hab, PNN50*, autonomic measures), visual reaction times and StartReact effect (*STR_Bic, VRT_Bic*, multimodal measures of sensorimotor function), and another measure of cortical excitability (*TMS_asymptote*). All bar one of these additional measures had individual significance levels P<0.05, but had failed the correction for multiple comparisons. This analysis suggests that there is little redundancy between measures and each captures a separate dimension of dysregulation in pCF.

## Discussion

The rapid development of successful vaccines against SARS-CoV-2 means that despite the evolution of variants, the majority of people in the UK are now largely protected from adverse short-term effects. Immunity also offers significant, but incomplete, protection from lingering sequelae^36^ and thus the incidence of pCF is likely to grow less rapidly than it has recently; nevertheless, the number still suffering remains staggering. Current estimates suggest ∼1% of the population have lasting fatigue, with enormous economic and social cost.

Much of our current research and understanding on the acute and chronic impacts of SARS-CoV-2 is centred on the inflammatory and immunological effects following an infection^37^, which in turn can affect many other systems in the body. Indeed, there is mounting evidence that inflammatory markers remain elevated several months after an infection for patients with the longer term sequelae^38^, but the relationship between inflammation and post-COVID fatigue remains unclear. Research from other chronic inflammatory conditions^39^ can be informative and shows that although fatigue is often associated with inflammation, a direct link between the two has proven elusive. Fatigue levels do not correlate well with circulating levels of inflammatory markers^40^. Many rheumatoid arthritis patients undergoing anti-inflammatory treatment still report high levels of fatigue, even though their disease itself is in remission^41^, suggesting that the relationship between fatigue and inflammation is not simple.

Although inflammation is likely to be important in the pathogenesis of pCF, a neural component is inevitable – the most common symptoms of pCF (as for fatigue in other conditions) are exhaustion after minimal physical or cognitive activity, both of which rely on neural circuits. There are multiple physiological pathways for the immune system to influence the nervous system and vice versa ^6^ but of particular interest is the fact that pro-inflammatory cytokines in the brain, that are elevated following an infection, can have profound effects on neuroplasticity^42 43^. Before being able to address whether such a mechanism operates in pCF, we first need to know which neural systems are affected.

In this study, we deployed an extensive battery of well-characterised non-invasive tests which are sensitive to different components of the nervous system. Although several measures were affected in pCF, it is important to emphasise that the majority of tests showed no difference between pCF sufferers and controls. Fatigue after SARS-CoV-2 infection does not result from a generalised deficit, but from specific changes in defined neural circuits. Our data support some of the hypotheses outlined in the Introduction, but also enable us to exclude some possible mechanisms.

Hypothesis 1 proposes that circuits providing inputs to motoneurons are less active in pCF; this could lead to weaker contractions, and an increased sense of effort. In support of this proposed mechanism intra-cortical facilitation, a measure of intracortical glutamatergic function^44^, was reduced in pCF^44^. Reduced facilitation was not countered by a concomitant reduction in intra-cortical GABAergic inhibition, suggesting a rebalancing of cortical activity and excitability to a lower level. As a result, corticospinal neurons could fire less vigorously for the same input from other upstream cortical areas, and hence plausibly lead to an increased sense of effort and fatigue. In agreement with these results, visual reaction times tended to be slower in pCF.

Hypothesis 2 suggests that fatigue results from an impairment of sensory attenuation during movement. If sense of effort is judged from the level of feedback, this could make a movement feel more effortful than it actually was, and hence lead to fatigue^7^. Importantly, a direct measure of sensory attenuation was unaffected in pCF; indeed, all measures which related to sensory processing appeared normal. While this mechanism may contribute to fatigue in other pathologies (e.g. after stroke, see ^10^), it does not appear important in pCF.

Hypothesis 3 is that pCF leads to myopathy, producing muscle weakness which requires an increased neural drive to generate a given contraction strength. Our results provide partial support for this idea. Individuals with pCF had normal grip strength, and there was no evidence of fatiguing transmission at the neuromuscular junction. As far as we could assess, the intrinsic excitability of motoneurons was also normal. However, myopathic changes became apparent after a sustained contraction, when the ability of muscle to produce force in response to electrical stimulation was significantly reduced in pCF subjects. This may reflect abnormalities in energy metabolism, leading to more rapid depletion of muscle energy stores^45^. Clearly such deficits could lead to a feeling of fatigue^46,47^, although whether muscle is regularly pushed into the regime where such effects become noticeable in everyday life is perhaps debatable.

Hypothesis 4 relates to the extensive role played by neuromodulators in motoneuron function^15^. Recent work has emphasised how active channels in the motoneuron dendrites amplify synaptic currents, and even generate sustained firing and thereby contractions in the absence of synaptic drive. The magnitude of these persistent inward currents is regulated by neuromodulators^48^. There is evidence for changes in neuromodulatory centres following other inflammatory^49^ or autoimmune disorders^16^; thus even a small reduction in tonic levels of neuromodulators could leave motoneurons relatively unresponsive to descending drive^50^, and hence generate feelings of weakness and fatigue. However, assessment of persistent inward currents showed no evidence for a difference in pCF, suggesting that this mechanism does not contribute to fatigue after SARS-CoV-2 infection.

Hypothesis 5 posits a role for the autonomic nervous system in fatigue^17,51,52^, and supporting this we found multiple abnormalities in autonomic function. Resting heart rate was elevated, and heart rate variability reduced; this suggests a rebalancing of parasympathetic *versus* sympathetic drive in favour of the latter. Habituation of the galvanic skin response to a loud (startling) sound was also reduced in pCF subjects, again supportive of excessive sympathetic output. Core body temperature was elevated, and blood oxygen saturation reduced. These metrics may reflect continued long-lasting impacts of the original infection on lung function and immune activation, but they may also result from a generalised heightened sympathetic tone.

An ongoing challenge with fatigue is to determine the extent to which it is caused by disordered physiology versus psychological and social factors. Blindly accepting all reported symptoms as having an organic origin, versus uniformly rejecting the lived experience of fatigue sufferers, are equally unsatisfactory clinical approaches. In this study, we were able to identify a small number of metrics with abnormalities in pCF. Using these alongside immunological biomarkers^38^ may allow a more objective diagnosis on the basis of signs rather than symptoms alone. Interestingly, there was no evidence for more than one cluster within the pCF cohort, as we might expect if pCF originated from multiple causes (which could include a psychogenic origin). This finding should be treated as preliminary, given the relatively small size of our cohort, but it does suggest that treatment of pCF may not require extensive stratification to be successful.

An important and unavoidable limitation of our work is its cross-sectional nature. Although it seems natural to assume that changes in metrics were caused by pCF, it is equally possible that these were present prior to the SAR-CoV-2 infection, and that these conferred increased risk for developing fatigue. We also do not know whether changes occurred early in the disease process prior to fatigue onset, or whether they developed alongside fatigue. These possibilities should be examined by a longitudinal study of individuals earlier after infection; objective metrics could help to identify individuals at risk of developing pCF, for whom more proactive management of an otherwise mild acute infection might then be warranted.

In conclusion, our results provide evidence of dysregulation in all three main divisions of the nervous system, using tests which are straightforward to administer and could easily be incorporated in future trials to assess and treat pCF. Knowledge of which neural circuits are involved in pCF may aid the development of novel, principled therapeutics. Whether these results are applicable to other post-viral fatigue syndromes as well as chronic fatigue remains to be determined.

## Data Availability

All data produced in the present study are available upon reasonable request to the authors; supplementary tables will be made available with the accepted manuscript.

## Abbreviations

Abbreviations in italics refer to specific measurements, which are outlined in Figure 1 and fully detailed in Supplementary Information.

ANS: Autonomic nervous system
*CMR_E1*: The early excitatory phase of the cutaneomuscular reflex
*CMR_E2*: The late excitatory phase of the cutaneomuscular reflex
*CMR_I1*: The early inhibitory phase of the cutaneomuscular reflex
COVID: Coronavirus disease
FIS: Fatigue impact scale
*Grip*: The maximum grip force
*GSR_Hab*: Habituation of the galvanic skin response following a loud sound
*Mean HR*: Mean heart rate
pCF: post-COVID fatigue
*pNN50*: Proportion of successive heart beat intervals which differ by more than 50 ms
PNS: Peripheral nervous system
*pO*_*2*_: Blood oxygen saturation
*RNS*: Repetitive nerve stimulation
SARS-CoV-2: Severe acute respiratory syndrome coronavirus 2
*STR_1DI*: The StartReact effect (speeding up of reaction time by a loud sound) measured in the first dorsal interosseous muscle
*STR_Bic*: The StartReact effect (speeding up of reaction time by a loud sound) measured in the biceps muscle
*SAT*: Sensory attenuation with movement
*SMU_AHP*: Estimate of the duration of motoneuron afterhyperpolarisation made from single motor unit discharge
*SMU_deltaF*: Difference in firing rate between recruitment and de-recruitment of single motor units
*SMU_peakF*: Peak firing rate in single motor units
*SMU_R2*: Correlation coefficient between smoothed discharge of single motor units
*SSRT*: Stop signal reaction time
*TDT*: Temporal difference threshold
*Temp*: Tympanic temperature
*TI_CA_baseline*: Central activation measured by twitch interpolation at baseline
*TI_CA_fatigued*: Central activation measured by twitch interpolation after a fatiguing contraction
*TI_PeriphFatigue*: Peripheral fatigue measured during twitch interpolation experiment
TMS: Transcranial magnetic brain stimulation
*TMS_AMT*: Active motor threshold of TMS
*TMS_asymptote*: The asymptote of the sigmoid fit to the curve of TMS response *vs* stimulus intensity
*TMS_CSP_AMT*: The cortical silent period duration after TMS at an intensity equal to the active motor threshold
*TMS_CSP_slope*: The slope of the relation between cortical silent period duration after TMS and stimulus intensity
*TMS_CSP_I50*: The cortical silent period duration after TMS at the intensity which generates a half-maximal response
*TMS_I50*: Intensity of TMS which generates a half-maximal response
*TMS_ICF*: Intracortical facilitation measured with TMS
*TMS_RMT*: Resting motor threshold of TMS
*TMS_SAI*: Short latency afferent inhibition measured with TMS
*TMS_SICI*: Intracortical inhibition measured with TMS
*TMS_slope*: Measure related to the slope of the sigmoid fit to the curve of TMS response *vs* stimulus intensity
*VRT_1DI*: Visual reaction time measured in the first dorsal interosseous muscle
*VRT_Bic*: Visual reaction time measured in the biceps muscle

## Acknowledgements

The authors wish to thank Norman Charlton for mechanical engineering support.

## Funding

This work was supported by a grant (MR/W004798/1) from the Medical Research Council (UKRI).

## Competing Interests

The authors report no competing interests.

## Supplementary Material

A detailed description of the methods, together with a spreadsheet listing Z-transformed measurements for each subject, is available in the Supplementary Material online.

## Notes

### Competing Interest Statement

The authors have declared no competing interest.

### Funding Statement

This study was funded by the Medical Research Council (MR/W004798/1, UKRI)

### Author Declarations

All studies were approved by the Ethics Committee of the Newcastle University Faculty of Medical Sciences; participants provided written informed consent to take part.

