## Supplementary Methods for "Neural Dysregulation in Post-COVID Fatigue"

### Supplementary Information

#### General Electrophysiological Methods

Electromyogram (EMG) was recorded with surface electrodes (Kendall H59P, Covidien, Dublin, Ireland) using an isolated amplifier (D360, Digitimer, Welwyn Garden City, UK; gain 500, bandpass 30 Hz-2 kHz). Where a measurement required a constant contraction, the amplitude of smoothed, rectified EMG was fed back to the subject via a display of coloured bars on a computer screen, calibrated to the individual’s maximum voluntary contraction (MVC). Subjects were asked to maintain these bars in the zone corresponding to 10% MVC. Stimuli to peripheral nerves (0.2 ms pulse width) were given with either a Digitimer DS7AH or DS5 isolated, constant current stimulator. Transcranial magnetic brain stimulation (TMS 0.2 Hz stimulus rate) was given with a Bistim 200^2^ stimulator and figure-of-eight coil (7 cm diameter of each winding; Magstim Company Limited, Whitland, UK), with the coil held tangential to the head at around 45° to the parasagittal plane, inducing current in the brain from posterior to anterior. Coil position relative to the head was maintained using a Brainsight neuronavigation system (Brainbox, Cardiff, UK). Stimulus timing was controlled by a Power1401 intelligent laboratory interface running Spike2 software (Cambridge Electronic Design, Cambridge, UK), which also sampled EMG and other task-related signals to hard disc (sampling rate 5 kSamples/s). All measurements were made on the self-reported dominant side. Offline analysis was performed with custom scripts written in the Matlab programming environment.

Thirty-three measures were chosen to provide non-invasive assessments of a wide range of cortical, brainstem and spinal neural circuits, as well as the function of the autonomic and peripheral nervous system. In addition, blood oxygen saturation and tympanic temperature were recorded, giving 35 measures in all. In the following, full details of each test and the measures derived from it are given. Measures are referred to in bold font with the same abbreviations used in Figs 1&2.

#### Repetitive Nerve Stimulation

This test measured the function of the neuromuscular junction. The median nerve was stimulated at the wrist, and EMG was recorded from the abductor pollicis brevis (AbPB) muscle. Stimulus intensity was set to produce a supra-maximal M wave. Trains of ten stimuli were given (3Hz), with ten repetitions (inter-train interval 10s). The peak-to-peak amplitude of the M wave elicited by the first and last stimulus in the train was measured from averaged traces. The size of the first response (M_max_) was used for subsequent normalisation of other measures (see below). The ratio of the last to the first response was calculated (repetitive nerve stimulation, measure *RNS*). In healthy subjects this is around unity; low values indicate a failure of neuromuscular transmission.

#### TMS Recruitment Curve and Cortical Silent Period

The increase in response with increasing stimulus intensity was used to measure motor cortical excitability^2^; the duration of the cortical silent period assessed intracortical inhibition^3,4^. EMG was recorded from the AbPB muscle. The TMS coil was moved to locate the hot spot for motor evoked potentials (MEPs). The active motor threshold (measure *TMS_AMT*) was determined. We then delivered sets of ten stimuli at AMT, and successive increments of 10% MSO, until 100% MSO, while the subject maintained an active contraction. Offline analysis measured the peak-to-peak of MEPs from single trials, and plotted this versus stimulus intensity. A sigmoid curve was fitted (using the MATLAB function *fminsearch*; fit repeated 200 times with random initial values and the best fit used) to the relationship^5^, according to:

$$MEP=\frac{{MEP}_{max}}{1+\exp\left( \frac{I_{50}-I}{k} \right)}$$

Where *MEP* is the MEP amplitude at intensity *I*, *MEP_max_* is the asymptote of the sigmoid relationship, *I_50_* is the intensity with a response of *MEP_max_/2* (measure *TMS_I50*), and k relates to the slope of the sigmoid curve (measure *TMS_slope*). *MEP_max_* was normalised as a percentage of *M_max_* to yield the measure *TMS_asymptote*.

Averages of rectified EMG were used to assess the duration of the cortical silent period, determined as the first time where the average returned to the pre-stimulus baseline level. Because silent period increases with stimulus intensity^6,7^, the duration was plotted against stimulus intensity, and a straight line fitted. The slope of this line yielded measure *TMS_CSP_slope***.** The estimated silent period duration at the active motor threshold gave measure *TMS_CSP_AMT*, and at the intensity *I_50_* gave measure *TMS_CSP_I50*.

#### Short-latency Afferent Inhibition

This test assessed the activation of cortical inhibitory circuits following stimulation of peripheral afferents^8^. EMG was recorded from the AbPB muscle, and the TMS coil was held over the same location as for the recruitment curve. The resting motor threshold (RMT) was determined, with an accuracy of 1% of maximum stimulator output (MSO), as the intensity which produced a MEP > 100 µV amplitude on 3/6 stimuli. TMS stimulus intensity was then set to produce a MEP peak-to-peak amplitude around 1mV, or to 1.2xRMT, whichever was lower. The median nerve was stimulated at the wrist, and the intensity adjusted to be at motor threshold, judged from the appearance of an M wave in the EMG. We then measured responses to TMS alone, and TMS preceded by median nerve stimulation at intervals of 20.8 to 25.8 ms in steps of 1ms. Twenty repetitions of each condition were given, in pseudo-random order, with the subject at rest. Offline analysis found the peak-to-peak amplitude of responses to TMS conditioned by nerve stimulation, as a percentage of TMS alone. The stimulus interval with greatest short-latency afferent inhibition yielded measure *TMS_SAI*.

#### Paired-Pulse TMS

This test assessed intra-cortical excitatory and inhibitory circuits. EMG was recorded from the first-dorsal interosseous (1DI); the TMS coil was moved to locate the hot spot for this muscle. The RMT was determined, as the intensity required to generate MEPs of amplitude greater than 100 µV on 3/6 sweeps, and used as measure *TMS_RMT*. The test stimulus intensity was set to generate an MEP amplitude of 1 mV, or to 1.2xRMT, whichever was lower. The conditioning stimulus intensity was 0.8xRMT. We then measured the responses to the test stimulus alone, and when preceded by the conditioning stimulus at intervals of 3 and 10 ms, corresponding to short-interval intracortical inhibition (SICI) and intracortical facilitation (ICF) respectively^9^. Twenty repetitions of each condition were given, in pseudo-random order, with the subject at rest. Offline analysis measured the peak-to-peak amplitude of the conditioning stimuli as a percentage of the responses to test stimulus alone, yielding measures *TMS_SICI* and *TMS_ICF*.

#### Stop-Signal Reaction Time

The stop-signal reaction time (SSRT) is a measure of response inhibition^10^. Here we followed a recently-developed modified procedure, which uses a portable device and Bayesian statistical analysis to improve the reliability of the measure^11^. Participants held a battery-powered microprocessor-controlled box, and pressed a button to initiate a trial. When a green LED illuminated, they were required to respond by releasing this button as quickly as possible. On 25% of trials, a red LED illuminated at 5, 65, 135 or 195 ms after the green LED; subjects were asked not to respond on these trials. Three blocks of 64 trials were recorded, with a 60 s break in between blocks; each block consisted of 48 Go trials, and 16 Stop trials (4 at each delay). The room lights were dimmed for this test. Using the distribution of reaction times on the Go trials, and the proportion of successfully inhibited responses, the algorithm calculated the SSRT as described in full in our previous work^11^, producing measure *SSRT*.

#### Temporal Difference Threshold

The temporal difference threshold (TDT) measures the ability to detect temporal offsets in timing of two stimuli, and is thought to reflect both cortical and subcortical function^12^. We implemented this measure using a portable box and Bayesian algorithm, similar to that used for SSRT. The TDT box contained two red LEDs and two response buttons. On each trial, the LEDs flashed for 1 ms, with a time separation between flash onset of *d* ms; participants were asked to press the left button if they saw simultaneous flashes, or the right button if they saw asynchronous flashes. The room lights were dimmed for this test. We assumed that the probability of the subject reporting asynchronous flashes followed a sigmoid curve:

$P\left( report Asynchronous with delay d \right|d_{50},k)=\frac{1}{1+exp\left( ((d_{50}-d)/k \right)}$

Where d_50_ is the separation at which subjects report asynchronous flashes 50% of the time, and *k* relates to the slope of the curve. The probability of the subject reporting a simultaneous flash is then simply:

$$P\left( report Simultanous | d_{50},k \right)=1-P\left( report Asynchronous | d_{50},k \right)$$

From Bayes’ rule:

$$P\left( d_{50},k \right|Subject response)=P\left( Subject response \right|d_{50}, k)\frac{P(d_{50},k)}{P(Subject response)}$$

The terms *P(d_50_,k)* and *P(Subject response)* correspond to priors in a Bayesian framework, here assumed uniform. This formula allows us to compute the probability of parameters *d_50_* and *k* assuming certain values, given the observed response of the subject. The parameter *d_50_* is the measure of interest in this test (corresponding to the TDT); the slope parameter *k* is a nuisance parameter, which may be removed by marginalisation:

$$P\left( d_{50} | Subject response \right)= \int_{k_{min}}^{k_{max}} P\left( d_{50},k \right|Subject response)dk$$

Where the limits of integration *k_min_* and *k_max_* were chosen based on prior expectation of slopes (here we used *k_min_*=0.6ms, *k_max_*=6ms).

The portable box implemented this algorithm iteratively in real time. Initially, a uniform prior distribution was assumed for *P(d_50_)*. Two trials were delivered, with *d*=0 and 120 ms, and the estimated probability distribution *P(d_50_|Subject response)* was then calculated. Two values of *d* to be tested next were determined as either the 1% and 50%, 50% and 75%, or 50% and 99% points of this distribution (which of these three options was used determined at random). This ensured that some ‘easy’ trials, which were clearly simultaneous/asynchronous, were mixed in with more ambiguous trials; we found this important to maintain subject motivation. The value of *P(d_50_|Subject response)* was then used as the prior *P(d_50_)* for the next two trials. The test continued until either 75 pairs of trials had been tested, or the interval between the 2.5% and 97.5% points of the *P(d_50_)* distribution (corresponding to the 95% confidence limits on the TDT estimate) was smaller than 3 ms. The box then reported the mean of the *P(d_50_)* distribution, which was used as the measure *TDT*.

#### Cutaneomuscular Reflex

The different components of the cutaneomuscular reflex assess the excitability of spinal and cortical excitatory and inhibitory circuits^13^. EMG was recorded from the 1DI muscle. Ring electrodes were placed on the middle and proximal phalanges of the index finger; stimulus intensity was increased gradually until just perceived by the subject (perceptual threshold, PT). Stimuli were then given at 3xPT, in ten blocks. Each block began with a brief sound cue, which instructed the subject to contract 1DI at 10% MVC. Two seconds after the beep, stimuli commenced (inter-stimulus interval chosen randomly 0.1250-0.1762 s, uniform distribution, n=100 stimuli). At the end of the stimulus block, the subject rested for 30 s before the next block, to avoid fatigue. Analysis used averages of rectified EMG. The amplitude of the E1, I1 and E2 components of the response^13^ were measured as the maximum (for E1/ E2) or minimum (I1) level above or below baseline, expressed as a percentage of the baseline, yielding measures *CMR_E1*, *CMR_I1* and *CMR_E2*. Reflex amplitudes were assessed from averages of all 1000 stimuli.

#### Sensory Attenuation with Movement

Sensory inputs are markedly attenuated during voluntary movement, due partly to descending control of feedback gain^14^. Deficits in this process have been previously hypothesised to be related to fatigue after stroke^15^. In this test, we obtained a quantitative estimate of sensory attenuation. EMG was recorded from the 1DI muscle. Stimuli were given to the digital nerves of the index finger, using adhesive surface electrodes placed on the proximal and middle phalanges. Subjects were required to report whether they detected a stimulus, in two conditions. The rest condition began when an automated voice cue ‘Rest Trial’ was played to the subject. A stimulus was then given (P=0.8) or not (P=0.2). A voice cue ‘Respond’ than asked the subject to report verbally if the stimulus was felt (yes/no). The movement condition began with a voice cue ‘Movement Trial’. The subject then made a rapid index finger abduction movement. The time of 1DI muscle EMG rising above a threshold was determined; the threshold was set to avoid noise triggers, but reliably to detect movements. Fifty milliseconds after this threshold crossing, a stimulus was given (P=0.8) or not (P=0.2). A voice cue ‘Respond’ then asked the subject to report detection (yes/no) as before. Rest and movement trials (n=50 of each) were given alternately. Stimulus intensity was decreased or increased for the next trial, depending on whether the stimulus was detected or not, with intensities for movement and rest trials being controlled independently.

Analysis consisted of fitting the probability of detection at intensity I to a sigmoid curve:

$$P\left( Detection at intensity I \right|I_{50},k)=\frac{1}{1+exp\left( ((I_{50}-I)/k \right)}$$

Where *I_50_* is the intensity with 50% detection, and *k* determines the slope of the curve. Catch trials, where no stimulus was given, typically had very low or zero detection probabilities, validating this model. The ratio of *I_50_* determined from movement to rest trials was used as the measure *SAT*.

#### Galvanic Skin Response Habituation

The galvanic skin response (GSR) is a change in the resistance of the skin generated by sweat production from sympathetic system activation^16^; its habituation may be a relevant measure in assessing cognitive states^17^. We measured the GSR by placing two metal plates on the lateral and medial surfaces of the index finger. With the subject sitting quietly at rest, five loud sounds were played through loudspeakers placed in front of the subject chair (115 dB, C weighting, 500 Hz, 50 ms, 6-6.8 s inter-stimulus interval, chosen randomly from a uniform distribution). The ratio of the GSR amplitude following the last stimulus compared to the first was used as measure *GSR_Hab*.

#### StartReact Effect

The StartReact effect is the shortening of voluntary reaction time by a loud (startling) sound; this has previously been used to assess connections from the reticulospinal system^18-20^. EMG was recorded from the 1DI and biceps muscles. Participants viewed a red LED, placed around 0.5 m in front of them. When this LED flashed (50 ms), they were instructed to perform an elbow flexion movement, combined with a clench of the fist, as quickly as possible. This generated a robust activation of the two EMG channels. A total of 60 trials were tested. For 20 trials, the LED flashed alone (visual reaction time, VRT); for 20 trials, the LED was combined with a quiet sound (81 dB, C weighting, 500 Hz, 50 ms; visual auditory reaction time, VART); for 20 trials, the LED was combined with a loud sound (115 dB, C weighting, 500 Hz, 50 ms; visual startle reaction time, VSRT). Trials were separated by 6-6.8 s, chosen randomly from a uniform distribution; the different trial types were delivered in random order. StartReact measurements were performed immediately after the GSR Habituation test, ensuring that any overt startle reflex had been habituated by the five loud sounds given in that test. The room lights were dimmed for this test. Offline analysis measured the reaction time on single trials as the point where EMG exceeded the baseline ± 7 SD; all trials were visually inspected, and automatically detected times corrected if they had resulted from noise or movement artifacts. Average VRT, VART and VSRT were calculated for each subject and muscle, together with the amplitude of the StartReact effect, equal to VSRT-VART. This yielded measures *VRT_1DI*, *VRT_Bic*, *STR_1DI* and *STR_Bic*.

#### Grip Force

Grip force is a well-validated measure of physical strength, which is reduced in conditions as varied as sarcopenia^21^ and cognitive decline^22^. Participants were seated, and held a hand grip dynamometer (model G200, Biometrics Ltd, Newport, UK) in their dominant hand, with the elbow flexed to 90° and the shoulder slightly abducted to position the dynamometer away from the body. They were asked to perform a maximal grip three times, with 60 s breaks in between. The largest force exerted over these three trials was taken as the grip strength, measure *Grip*.

#### Twitch Interpolation

The twitch interpolation (TI) procedure allows assessment of an individual’s ability to activate muscle maximally; in this study, we also measured changes after a sustained (fatiguing) contraction^23^. The protocol followed previous work from this laboratory^24^. Subjects sat with their arm and forearm strapped into a dynamometer to measure torque about the elbow; the shoulder was flexed, and the elbow at a right angle, so that the upper arm was horizontal and the forearm vertical. The forearm was supinated. Thin stainless-steel plate electrodes (size 30x15 mm) were wrapped in saline-soaked cotton gauze and taped over the belly of the biceps muscle and its distal tendon. Electrical stimuli were delivered through these electrodes while monitoring the evoked twitch response recorded by the dynamometer, and the intensity increased until the response grew no further. This supramaximal stimulus was used for all subsequent measurements.

The following recordings were then made in sequence. A brief tone cued the subject to make and hold a maximal voluntary contraction; 2 s after the tone, a stimulus was given to the biceps. After 1 s, a second tone indicated that the subject should relax. Five seconds later, a further biceps stimulus was given, followed by a further 55 s rest period. This sequence was repeated three times. A long tone then cued the subject to make a sustained maximal voluntary contraction. This was continued either for 90 s, or until the force exerted fell to 60% of the initial maximal level. During this sustained contraction, the biceps was stimulated every 10 s. After the contraction ended, a final three biceps stimuli were given at rest (inter-stimulus interval 5 s).

Averages of twitch response were compiled from these stimuli, and the force at the peak of the twitch relative to the pre-stimulus baseline measured. From the three stimuli delivered at rest at the start, we measured the maximal twitch at rest, $F_{rest}^{before}$. From the three stimuli delivered during MVC at the start, we measured the maximal twitch during contraction, $F_{MVC}^{before}$. From the final three stimuli delivered during the sustained contraction, we measured $F_{MVC}^{after}$. From the three stimuli delivered at rest after the sustained contraction, we measured $F_{rest}^{after}$.

If a subject truly performs a maximal voluntary contraction, a superimposed electrical stimulus should not be capable of generating extra force. The size of any elicited twitch thus measures a central activation deficit. Accordingly, we calculated central activation before fatigue (measure *TI_CA_baseline*) as:

$$TI\text{\_}CA\text{\_}baseline=\left( 1-\frac{F_{MVC}^{before}}{F_{rest}^{before}} \right)100\%$$

Central activation after fatigue (measure *TI_CA_fatigued*) was likewise calculated as:

$$TI\text{\_}CA\text{\_}fatigued=\left( 1-\frac{F_{MVC}^{after}}{F_{rest}^{after}} \right)100\%$$

Peripheral fatigue (measure *TI_PeriphFatigue*) was calculated as:

$$TI\text{\_}PeriphFatigue=\frac{F_{rest}^{after}}{F_{rest}^{before}} 100\%$$

This describes the reduced ability of the muscle to generate force after fatigue, even when activation is performed independent of the central nervous system by an electrical stimulus to the muscle.

#### Heart Rate and Heart Rate Variability

Heart rate and its variability can provide important insights into autonomic function^25,26^. A single channel ECG recording was made, using a differential recording from either left shoulder and right leg, or left and right shoulders (bandpass 0.3-30 Hz, gain 500). The ECG was processed offline to extract the time of each QRS complex. From these times, we computed the mean heart rate (measure *Mean HR*), and the *pNN50*. This is a measure of heart rate variability^27^ defined as the proportion of successive intervals which differ by >50 ms. Heart rate measures were made during the SSRT test (see above), which ensured that the subject was sitting quietly, while engaged in a consistent behaviour.

#### Motoneuron Physiology

Motoneurons have active channels, which can amplify and modulate responses to synaptic input^28^. This test was designed to derive measures of motoneuron function, using high density surface EMG recordings to extract the activity of single motor units. Participants sat in a chair, with the leg on the dominant side outstretched and the knee straight. The foot was strapped into a rigid device which resisted movement around the ankle joint; the leg was also strapped down. A surface EMG grid electrode (13x5 electrodes, 8 mm inter-electrode spacing, part number GR08MM1305, OT Bioelettronica, Turin, Italy) was placed over the tibialis anterior muscle, and connected to a custom preamplifier (based on RHD2164 integrated circuit and RHD 512 channel recording system, Intan Technologies, Los Angeles, USA; bandwidth 10 Hz – 5 kHz, sampling rate 10 kSamples/s). Reference and ground electrodes were standard adhesive electrodes as used in other tests, placed on the patella and nearby skin respectively. The output of a single channel of EMG data was routed in real time to the Power1401 system and Spike2 software used for all other recordings. A custom script within this system presented subjects with a desired triangular activity profile, comprising 5 s rest, a linear increase to 30% MVC over 10 s, a linear decrease to rest over 10 s, and 5 s rest. At the start of a trial (signalled by an auditory cue), an overlain line was displayed on the desired profile, corresponding to the smoothed rectified EMG; subjects were instructed to track the target profile as closely as possible and to avoid sudden trajectory corrections. We measured a total of 15 trials, separated by 30 s rests to avoid fatigue. Finally, the subject performed a further 4 trials, with steady contractions at 10% MVC for 20 s.

#### Motor unit Decomposition and Analyses

The signals from the high density surface EMG grid over the TA muscle were decomposed into motor unit spike trains with a blind source separation algorithm^29^. The motor unit spike trains were visually inspected and corrected by experienced examiners, according to the guidelines described elsewhere^30^. Motor units with high inter-spike variability (i.e., mean coefficient of variation above 40% and a silhouette measure below 0.92)^29^ were discarded since they are typically associated with intermittent activation. The extracted MU spike trains were subsequently used to estimate several parameters relating to motoneuronal physiology.

Delta F: We used paired motor unit analysis to quantify the level of hysteresis (also known as ΔF) in the recruitment and derecruitment of the recorded MUs during the ramp contractions only, as this can provide a measure of persistent inward currents and the neuromodulatory drive to the muscle. ΔF was calculated as the difference in the activity of a lower threshold ‘control’ MU at the times of recruitment and derecruitment of a higher threshold MU. MU firing is variable and non-stationary during the ramp; to smooth the activity profile a third order polynomial was fitted for each MU and rate and onset measurements were taken from smoothed firing rates. We excluded ramp trials with sudden changes in muscle activity, where the EMG gradient during the ascending and descending phases of the ramp were significantly different. This was determined by fitting a linear regression line in the rectified TA EMG through the ascending and descending ramp phases; if the 95% CI of the gradient overlapped, they were assumed not to be different. Using the criteria specified^31^ for the selection of suitable pairs of MUs, we extracted multiple ΔF measures for each subject - the median of those values was selected as representative across the TA motor pool, and used as measure *SMU_deltaF*.

Peak F: For each decomposed motor unit, the maximum value of the smoothed instantaneous firing rate profile was taken across all ramps that the motor unit was decomposed for. The median value across all MUs for a given subject was selected as representative for the TA motor pool for that subject, and gave measure *SMU_peakF*.

After-Hyperpolarisation potential estimate (AHP): The inter-spike interval histogram of a spike train can be transformed to provide a measure of the time course of the cell membrane trajectory after a spike, and this can be used to infer several of the physiological properties of the cell. Of particular interest is the AHP duration: this is correlated with the motor unit twitch time^32^, so that motoneurons with long AHPs tend to innervate slow-contracting motor units^33^. Differences in the AHP duration between pCF and control could suggest changes in the properties of motoneurons.

AHP trajectories were extracted following the procedure described in detail previously^34-36^. This began with the inter-spike interval histogram (1ms bin width), from which was calculated the death rate - this is the probability that an interval will end at a given time delay after the previous spike. The death rate profile was then converted to a distance to threshold trajectory by using a random walk model of a neuron responding to noisy input. Distance to threshold trajectories were formed for inter-spike intervals selected, on the basis of adjoining intervals, to come from a period of homogenous firing rate; these trajectories were then combined to generate a compound AHP trajectory. Once the shape of the AHP was calculated, it was fitted with a first order exponential curve. In most cases the Spearman’s correlation coefficient between the AHP and the fitted exponential was > 0.9; units were excluded if they had correlation values lower than this. From the fitted exponential the time constant of the AHP could be measured. This process was applied to the spiking data collected during the ramp contractions. For each subject the median time constant of the AHP across all available units was used as measure *SMU_AHP*.

Common input measurement: Motoneurons show synchrony in moment-by-moment fluctuations in firing because they receive common inputs. To estimate the strength of common input across the pool of TA units, we measured the time-domain cross correlation between units in the pool during the steady contraction period^37^. The spike trains were randomly divided into two equally sized groups, summed and convolved with a 25ms Hanning window; the cross-correlation between the two spike trains was then estimated. This was repeated 25 times and the cross-correlation strength (R) for the pool of units was taken as the maximum value of the average cross-correlogram across the iterations. Any units with mean rates < 5Hz during the hold period were excluded from this analysis. The square of the cross-correlation was used as measure *SMU_R^2^*.

#### Biometric Data

In addition to the neural and behavioural measures, we also made biometric measurements. These included blood oxygen saturation (*pO_2_*), tympanic temperature (*Temp*), height and weight; the latter two measures were used to calculate the body mass index (measure *BMI*)^38^.

#### Multivariate Classification

Although we found a significant difference in several individual measures between the fatigue and control cohorts, the redundancy between such high dimensional data can be difficult to measure. To ascertain how useful our high dimensional neurophysiological features were in distinguishing between the two cohorts, we carried out linear classification analysis (routine *fitclinear* in MATLAB). We utilised the least absolute shrinkage and selection operator (lasso) for regularising our dataset, and to decide the best features to use for classification and hence avoid overfitting our model. The strength of the regularisation (or how strict the classifier was) was specified by the ‘lambda’ parameter. This analysis used normalised Z scores as described above.

We first estimated the most common number of features required for classification. To do this we implemented 5000 iterations of the classifier; each run used a different random choice to replace any missing data values (see section below). For each iteration, 40 logarithmically spaced lambda values were used, and cross-validation was carried out through multiple folds (parameter kfold=10 in the *fitclinear* routine). The average classification accuracy for each value of lambda across all the folds was obtained via the kfoldLoss method, and the lambda with the highest classification accuracy was selected for that iteration. For the chosen lambda value, the number of ‘useful’ features was counted as those whose weights were not reduced to zero. Across the 5000 iterations we compiled a histogram of the number of features (Fig. 2C) and chose the modal value, which for our dataset was 6. We then determined which features made the strongest contributions to the classification by retraining a classifier (5000 iterations) but with the number of significant features locked to 6. This was done (for each iteration) by adjusting the value of lambda so that 6 features had non-zero weights - the identity and weight of the surviving features was noted. The fraction of times that a given measure survived this selection process is shown in Fig. 2D.

#### Cluster Analysis

Although our two cohorts were significantly different in multiple measures, there were many possible schemata for the distribution of the measures within the pCF cohort - it could be that the significant differences were concentrated to a subset of the pCF participants, or that each measure was different only in a small separate sub-group of participants. To tease these possibilities apart we carried out clustering analysis (K-means clustering) in order to find the optimal number of clusters that would fit our dataset. For a given number of pre-defined clusters, k-means clustering initially chooses cluster centres randomly and data points are assigned to their closest cluster, based on their Euclidian distance from each cluster centroid. By then taking the mean of all data points within each cluster, a new set of clusters centres is determined and the process is repeated until the cluster centres stop moving between iterations. K-means clustering however cannot determine the optimal number of clusters. To do this we used the Gap Evaluation criterion^41^ which uses the biggest change in within-cluster distance between different cluster sizes to determine the optimal number of clusters. We tested for cluster numbers from 1 to 10.

We ran 100 iterations and for each iteration we extracted the Gap Distance for each cluster size (using the *evalcluster* function in MATLAB, with the ‘kmeans’ and ‘gap’ options selected). The mean and standard error across these iterations was plotted in Fig. 2B for each cluster number. We ran the cluster analysis by using only the four metrics that were most significant from our dataset, but also by using all of our measures. In both cases (Fig. 2B) the change in Gap value was largest in going from 1 to 2 clusters, suggesting that a cluster size of 1 is optimal, and that the metric differences were homogeneously distributed across the pCF cohort in this study.

#### Missing Data

All participants underwent the same battery of tests, but it was not possible to obtain all measures in all subjects (these are shown as blank spaces in Supplementary Table 1). For example, the measures of motoneuron physiology rely on the decomposition of sufficient numbers of motor units; this is not always possible, especially in subjects with substantial subcutaneous fat. For the paired analyses between pCF and control cohorts for individual metrics, missing values were excluded prior to comparison. For the classification analysis this was not possible, as it would severely limit the size of the cohorts. There are various approaches for imputing missing data^42^, but most rely on using values from other available features for a given subject to predict the missing datum. In this case, because we wished to make comparison between the different features, this approach could compromise our classification. Instead, for each cohort, missing data from a given feature was filled in by randomly selecting data from the subjects within that cohort for whom the data were available.

### Supplementary Tables

**Supplementary Table 1. Participant information and values of the 35 measures, for the two cohorts, pCF and control.** This table shows the biometric data, FIS scores, and neurophysiological measurements for each individual participant – neurophysiological values have been Z-normalised (as described above). Each row corresponds to a single participant and each column corresponds to a specific measure.

**Supplementary Table 2. Normalised and statistical test values for the 35 measures sampled in this study, for each cohort pCF and control.** This table shows the summary values for each metric we measured for the two cohorts separately. The mean, standard deviation and standard error of the mean are shown for each cohort, and for the pCF cohort we also show the Z-normalised values (relative to the control cohort). The column before last shows the statistical significance of an unpaired – test for each metric (with coloured rows indicating p<0.05). The final column shows the p-values adjusted for multiple comparisons (using the Benjamini-Hochberg approach described above) and rows with an asterisk (*) indicate those that cross the significance threshold adjusted for multiple comparisons.
